# Sensitivity of nasopharyngeal swabs and saliva for the detection of severe acute respiratory syndrome coronavirus 2 (SARS-CoV-2)

**DOI:** 10.1101/2020.05.01.20081026

**Authors:** Alainna J. Jamal, Mohammad Mozafarihashjin, Eric Coomes, Jeff Powis, Angel Xin Liu, Aimee Paterson, Sofia Anceva-Sami, Shiva Barati, Gloria Crowl, Amna Faheem, Lubna Farooqi, Saman Khan, Karren Prost, Susan Poutanen, Lily Yip, Zoe Zhong, Allison J. McGeer, Samira Mubareka, for the Toronto Invasive Bacterial Diseases Network COVID-19 Investigators

## Abstract

We enrolled 53 consecutive in-patients with COVID-19 at six hospitals in Toronto, Canada, and tested one nasopharyngeal swab/saliva sample pair from each patient for SARS-CoV-2. Overall, sensitivity was 89% for nasopharyngeal swabs and 77% for saliva (p=NS); difference in sensitivity was greatest for sample pairs collected later in illness.

## Background

Rapid and accurate detection of severe acute respiratory syndrome coronavirus 2 (SARS-CoV-2) in patient specimens is critical to controlling the coronavirus disease 2019 (COVID-19) pandemic. As yet, there are few data comparing sensitivity of different specimen types for SARS-CoV-2 detection.

In Canada, nasopharyngeal (NP) swabs are the preferred collection site for SARS-CoV-2 testing [1,2], and preliminary data suggest that they may be more sensitive than oropharyngeal swabs for SARS-CoV-2 detection [3,4]. However, collection of both NP and oropharyngeal swabs is uncomfortable for patients and may pose risk to healthcare workers. Moreover, recent global supply chain shortages have resulted in limited access to various swabs types. Saliva, in contrast, can be easily self-collected by adolescents and adults. Other groups have demonstrated successful detection of SARS-CoV-2 in saliva specimens and use of saliva for serial sampling [5-7]. We aimed to compare sensitivity of NP swabs and saliva for SARS-CoV-2 detection in hospitalized patients.

## Methods

The Toronto Invasive Bacterial Disease Network (TIBDN) performs population-based surveillance for select infectious diseases in metropolitan Toronto and the regional municipality of Peel (population base 4.2 million in 2016), Ontario, Canada. For COVID-19, clinical microbiology laboratories report specimens testing positive for SARS-CoV-2 to the central study office. Starting on March 16^th^, 2020, study staff enrolled consecutive in-patients at 6 TIBDN hospitals. Patient demographic, exposure, and medical data were collected by interview and chart review. An NP swab and saliva specimen were collected on the day of enrolment, and then 3 subsequent pairs of samples were obtained at 72-hour intervals if the patient remained hospitalized. NP swabs were collected as per standard procedures [8]. For saliva specimens, patients were asked to spit into a sterile specimen container and then 2.5 mL of phosphate-buffered saline were added.

Samples were transported to the research microbiology laboratory, where they were aliquoted and frozen at −80°C within 8 hours of collection. On April 14^th^, we selected each patient’s most recent NP swab/saliva sample pair for SARS-CoV-2 real-time polymerase chain reaction (RT-PCR) testing. Laboratory testing was with the Allplex™ 2019-nCoV Assay(100T) to detect RNA-dependent RNA polymerase (RdRp), envelope (E), and nucleocapsid (N) genes (Seegene Inc, Seoul, South Korea) at Sinai Health System (Toronto, Canada).

This study was approved by the Research Ethics Board of Sinai Health System (#02-0118-U).

## Results

Fifty-three in-patients were included; all were confirmed to have COVID-19 with an NP swab tested in a clinical laboratory in Toronto. The median age was 63 years (range 27-106), 21 (40%) were female, 38 (72%) had at least one comorbidity, and 4 (8%) were immunocompromised. Seven (13%) had a household contact as the suspected source of exposure. On admission, 47 (89%) had fever and 44 (83%) had cough. The median time from illness onset to hospital admission was 6 days (interquartile range (IQR) 3-8) and 18 (34%) required intensive care. The median time from illness onset to collection of the tested specimens was 11 days (IQR 7-15). As of April 22^nd^, 8 (15%) remained hospitalized, 41 (77%) were discharged, and 4 (8%) had died.

Of 53 patients with paired specimens tested, 47 (89%) had at least one positive specimen. In 31 (66%) of these 47 patients, both NP swab and saliva were positive, in 11 (23%) only the NP swab was positive, and in 5 (11%) only saliva was positive, p=0.14 (Table). Thus, using NP swabs only would have detected 42/47 (89%) patients with at least one positive specimen and using saliva only would have detected 36/47 (77%) patients with at least one positive specimen. Using NP swabs only would have detected 13/14 (93%), 18/22 (82%), and 11/11 (100%) patients in their first, second, and third/fourth week of illness, respectively (Table). Using saliva only would have detected 12/14 (86%), 17/22 (77%), and 7/11 (64%) patients in their first, second, and third/fourth week of illness, respectively (Table).

**Table.** Results of testing of NP swab and saliva for SARS-CoV-2 RNA in hospitalized patients with COVID-19, by time from illness onset to specimen collection (N=53).

|  | **No. (%) patients**<br>**with NP swabs**<br>**and saliva both**<br>**positive** | **No. (%)**<br>**patients with**<br>**NP swab only**<br>**positive** | **No. (%)**<br>**patients**<br>**with saliva**<br>**only positive** | **No (%)**<br>**patients with**<br>**NP swab and**<br>**saliva both**<br>**negative** |
| --- | --- | --- | --- | --- |
| **Time from illness onset**<br>**to specimen collection** |  |  |  |  |
| 0 to 7 days (n=14) | 11 (79) | 2 (14) | 1 (7) | 0 (0) |
| 8 to 14 days (n=24) | 13 (54) | 5 (21) | 4 (17) | 2 (8) |
| 15 to 26 days (n=15) | 7 (47) | 4 (27) | 0 (0) | 4 (27) |
| Any (0 to 26 days)<br>(n=53) | 31 (58) | 11 (21) | 5 (9) | 6 (11) |

The median N gene cycle threshold (Ct) for NP swabs was 33 (IQR 30-36) when the saliva specimen in the pair was positive (n=31) versus 32 (IQR 28-35) when the saliva specimen in the pair was negative (n=11) (p=0.1). Among the 31 paired NP swab/saliva specimens that were both positive, N gene Ct values in NP swabs and saliva were similar (median 32, IQR 28-35 and median 31, IQR 28-36, p=0.6, respectively); the Pearson correlation coefficient was 0.4, p=0.03 (Figure). Results were similar when Ct values of the E and RdRp genes were used (data not shown).

**Figure.**
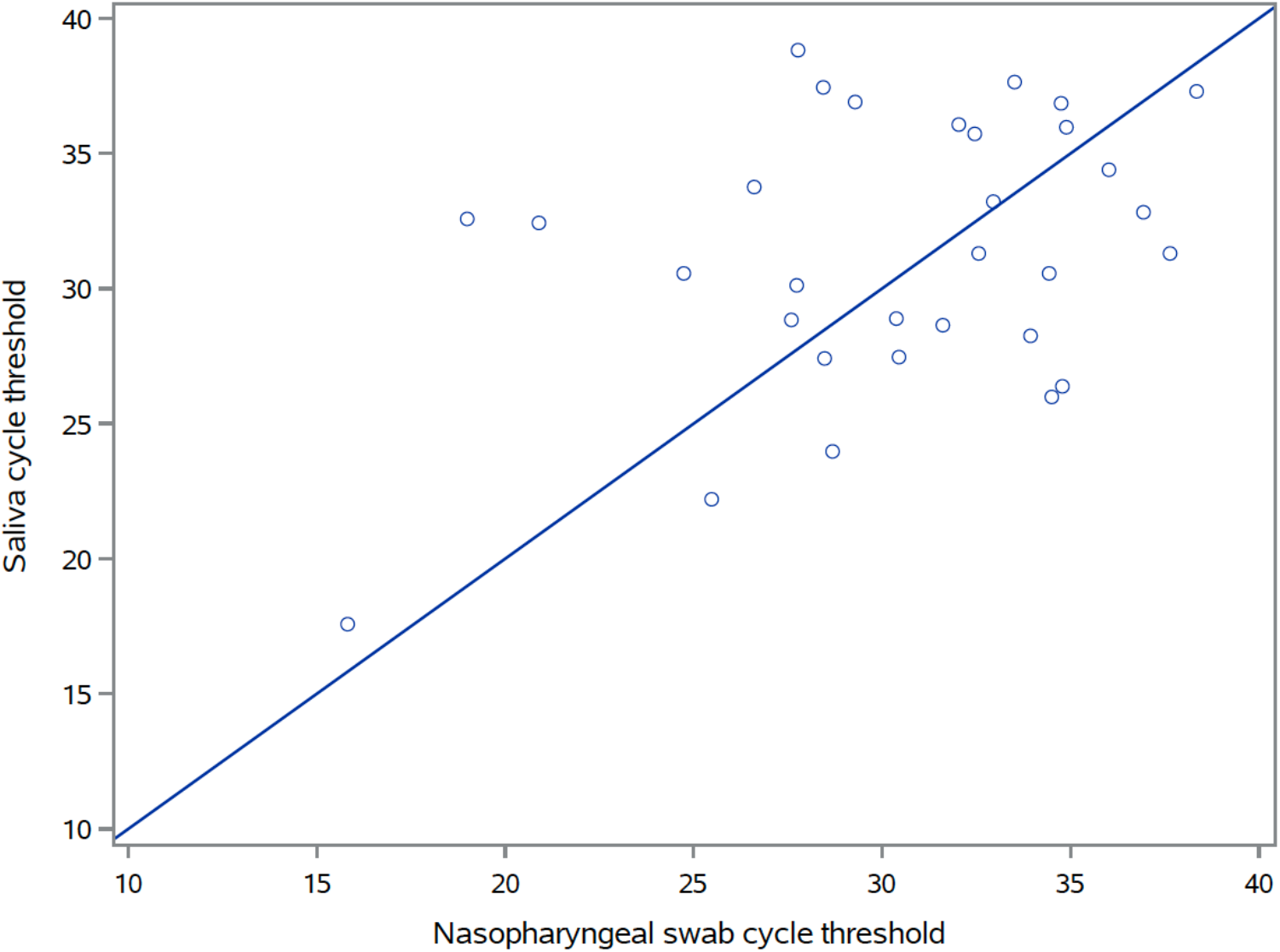
Nucleocapsid (N) gene cycle thresholds for nasopharyngeal swabs and saliva in pairs where both were positive for SARS-CoV-2 RNA (N=31).

## Discussion

In this sample of 53 in-patients, there was no significant difference in yield from NP swabs and saliva. However, NP swabs were approximately 10% more sensitive than saliva overall. The difference in sensitivity between NP swabs and saliva appeared to be greater if specimens were collected later in illness when viral concentrations have been reported to be lower [4,9]. Our data suggest that collection of NP swabs for SARS-CoV-2 detection in patients may be preferred, especially if the patient is later in illness.

Our data also suggest that neither a single NP swab nor a single saliva specimen is 100% sensitive for the detection of COVID-19. This is consistent with prior literature [10], emphasizing that a single negative test does not rule out disease in patients with a high pre-test probability of COVID-19. Repeated samples, or possibly serologic testing (particularly if the patient is in the second week of illness or later) [4], may improve yield.

There are several limitations to this analysis. As these patients were originally diagnosed using NP swabs, it is possible that there is a bias towards subsequent NP swabs versus other specimens being positive. We used a single detection system (Seegene), and other platforms may have yielded different results. We simply asked patients to spit into a specimen container; it is unclear whether other methods, such as throat washing [11], would have improved yield.

Our data suggest that saliva may be reasonably substituted for NP swabs in hospitalized patients when NP swabs are in short supply or patients cannot tolerate them, particularly early in illness when viral concentrations in the upper respiratory tract may be higher. However, this should be undertaken with the understanding that a single specimen is not 100% sensitive, and an NP swab should be used as a second specimen in patients for whom there is a high index of clinical suspicion and saliva is negative. More data are needed to determine whether saliva and NP swabs are truly equivalent early in illness, to assess testing on different platforms, and to assess the sensitivity of different specimen types in asymptomatic patients or those whose illness does not require hospitalization.

## Data Availability

Data are with the authors.

## Acknowledgements

We are grateful to the patients with COVID-19 who have agreed in difficult circumstances to help with research, and to the many laboratory, infection prevention and control, public health and research ethics staff who make the work of the Toronto Invasive Bacterial Diseases Network possible.

